# Making Broad Evidence Synthesis Feasible: An LLM Screening Agent for Meta-Analyses Applied To Suicide Prevention

**DOI:** 10.64898/2026.08.12.26360335

**Authors:** Daniel Dobin, Ashley M. Witmer, Fiona Gail Sweeney, Taylor Ryan, Andrea Cimino, Emily E. Haroz, Paul S. Nestadt, Holly C. Wilcox

## Abstract

**IMPORTANCE:** Systematic reviews and meta-analyses inform suicide-prevention policy and practice, but broad database searches are difficult to screen manually. This limits capture of upstream interventions, such as economic policies, with indirect effects on suicide. Reliable automated screening could make broader and more comprehensive evidence syntheses feasible.

**OBJECTIVE:** To develop and validate ScreenAgent, a large language model (LLM) agent for title and abstract screening, and a review-specific method for prospectively estimating screening performance.

**DESIGN, SETTING, AND PARTICIPANTS:** ScreenAgent was validated internally on a prospective meta-analysis, and externally on two published systematic reviews. The correct include and exclude decisions followed standard systematic-review screening methodology.

**EXPOSURES:** ScreenAgent, an LLM agent returning structured include-or-exclude decisions. Records it marked for inclusion were re-checked by a second, cascade pass using a higher-effort LLM. For the external reviews, the agent’s prompt was tuned automatically on a small set of labeled examples.

**MAIN OUTCOMES AND MEASURES:** We calculated sensitivity, specificity, workload reduction (the percentage of records removed from human review), and agent-versus-human reliability via Cohen kappa. Sensitivity was estimated by direct comparison (internal) and 5-fold cross-validation (external).

**RESULTS:** In the internal validation, ScreenAgent identified 43 of 44 eligible studies (sensitivity 97.7%; 95% CI, 88.2%-99.6%) with a generic prompt applied without any review-specific optimization, specificity 98.0%, and a measured full-corpus workload reduction of 99.4%. The cost was $855.91 for the full 201,064-record corpus (0.43 US cents per record). Agent-versus-human-consensus agreement exceeded human-versus-human agreement (Cohen kappa 0.75 vs 0.64; percent agreement 97.3% vs 95.4%). For two external validation studies, automatic tuning resulted in a cross-validated sensitivity of 95.9% (95% CI, 90.0%-98.4%) and 97.4% (90.9%-99.3%), with workload reductions of 97.4% and 98.4%.

**CONCLUSIONS AND RELEVANCE:** Suicide prevention efforts often require rapid consolidation of evidence because of the inherent challenges of single studies trying to prevent rare outcomes. On both internal and external validation sets, ScreenAgent identified nearly all eligible studies with human-level reliability for a fraction of a US cent per record while keeping human reviewers as the final arbiters. By making broad searches feasible and screening performance measurable beforehand, this approach can serve as a transparent methodology to strengthen the speed at which we can inform and advance suicide prevention efforts.

**Key point:** *Question:* Can an LLM agent screen records accurately to make broad-scale meta-analyses in suicide prevention more feasible?

*Findings:* Across a prospective review and two published reviews, ScreenAgent identified nearly all eligible studies (internal sensitivity 97.7%; external 95.9/97.4%) with human-level reliability (Cohen kappa 0.75 agent-vs-consensus, vs 0.64 between humans) and 98.0% specificity.

*Meaning:* Systematic reviews and meta-analyses are critical for evidence synthesis but are time and labor intensive. An LLM screening agent identified nearly all eligible studies with human-level reliability while reducing human workload by 99% at a fraction of a cent per record, which makes large-scale syntheses tractable.

## Introduction

There are 720,000 suicides worldwide each year and it is the third leading cause of death among those aged 15 to 29 years worldwide and second in the United States.^1,2^ Reducing suicide is a global and national priority: the 2024 National Strategy for Suicide Prevention names community-based suicide prevention as the first of its four strategic directions.^3^ Policymakers, public health officials, and clinicians rely on systematic reviews and meta-analyses to determine which programs, policies, and laws to implement.^4-7^ The validity of these syntheses heavily depends on whether their search and screening procedures identify the full range of relevant studies. Existing reviews may miss an important class of interventions: upstream programs and policies, such as those addressing social, structural, and economic risks, that reduce suicide deaths or attempts even though suicide prevention was not their primary purpose.^8^ Suicide risk is influenced by both community-based programs designed for suicide prevention (e.g., gatekeeper training, crisis hotlines)^9,10^ and interventions not primarily intended for suicide prevention (e.g., firearm and alcohol policies, healthcare insurance expansion).^11-13^ Policies and interventions designed to prevent other outcomes such as homicide, substance abuse, and violence as well as enhance coping skills and behavioral and emotional regulation share etiological factors with suicide can also prevent suicide-related outcomes.^14,15^ The efforts of interventions not primarily designed for suicide prevention but with effects on suicide-related outcomes are said to have “cross-over effects.” A fully comprehensive systematic meta-analysis would have to evaluate all of these efforts together, which is an existing gap hamstringing suicide prevention efforts at the local and global level.

Capturing crossover-effect studies presents a fundamental search problem. Studies of interventions explicitly designed for suicide prevention are indexed using terms like “suicide prevention,” and can therefore be identified through relatively focused searches.^4,5^ In contrast, studies of interventions developed for another purpose may report suicide only as a secondary outcome and would necessitate a much broader search term like “suicide.” Broad searches are more comprehensive, but may return hundreds of thousands of records, far more than a human team can feasibly screen.^16,17^ Researchers may therefore rely on narrow, highly specific search strategies to control workload. Although practical, this approach risks excluding relevant crossover-effect studies and limits the comprehensiveness of suicide prevention evidence synthesis. Automated screening offers a potential solution. Frontier large language models (LLMs) can read and classify text at a scale and speed not previously possible, raising the prospect of screening very large corpora without a restrictive search. Interest in this application has grown rapidly. A 2026 systematic review (not specific to suicide) identified 63 studies evaluating LLMs for systematic review tasks, comprising 148 performance assessments, title-and-abstract screening being the most common task.^18^ Earlier active-learning tools reduce workload but rely on iterative human labeling,^19-22^ and most LLM validations are retrospective and single-review, repeatedly showing high sensitivity undercut by low precision and unstable performance in the low-prevalence settings typical of broad searches.^23-26^ Early prospective and living-review deployments have begun to appear,^27-29^ but none target the broad, cross-domain searches needed to capture crossover-effect studies or ground screening performance in confidence intervals from a small human-labeled reference set.

We developed and validated ScreenAgent, an LLM agent for title-and-abstract screening specific to evidence synthesis for suicide prevention, and report two contributions. First, we apply automated screening to the broad, cross-domain searches required to capture crossover-effect studies, internally validating the agent on a prospective review (the Community-Based Suicide Prevention Synthesis) and testing it on two published external reviews to measure external validity. Second, we propose a methodology that validates the agent separately for each review in which it is used: human reviewers screen a small reference set in the usual way, that set is used to optimize and validate ScreenAgent’s prompt, and its sensitivity and specificity are estimated on it before the prompt is applied to the full, broad-range corpus. To our knowledge, no previous study has combined review-specific prospective validation with automated screening of crossover-effect studies in suicide prevention.

## Methods

### Study Design and Oversight

We developed and validated ScreenAgent, a large language model (LLM) agent for title-and-abstract screening in systematic reviews, in two stages. First, we conducted an internal validation within the Community-Based Suicide Prevention Synthesis (CBSPS) review, the prospective application study for which ScreenAgent was developed. Second, we assessed external validity against two recently published fully replicable systematic reviews on suicide outcomes with known eligibility decisions: Robinson et al., (2018) and Witt et al., (2021).^6,30^

For each record, ScreenAgent received the title, the abstract, and the eligibility criteria and returned a structured output consisting of a set of Boolean exclusion indicators, resulting in an include-or-exclude decision (Figure 1). The detailed agent specification is provided in eMethods 1. The primary outcomes were sensitivity and workload reduction (WLR). Sensitivity was prioritized because incorrectly excluding an eligible study was considered the more consequential error. Workload reduction was the percentage of records excluded by ScreenAgent, eliminating the need for downstream human screening. The study used published literature and the investigators’ own adjudicated screening decisions and did not involve any other human participants; institutional review board approval was therefore not required. The study follows the TRIPOD-LLM reporting guideline (eTable 9).^31^

**Figure 1.**
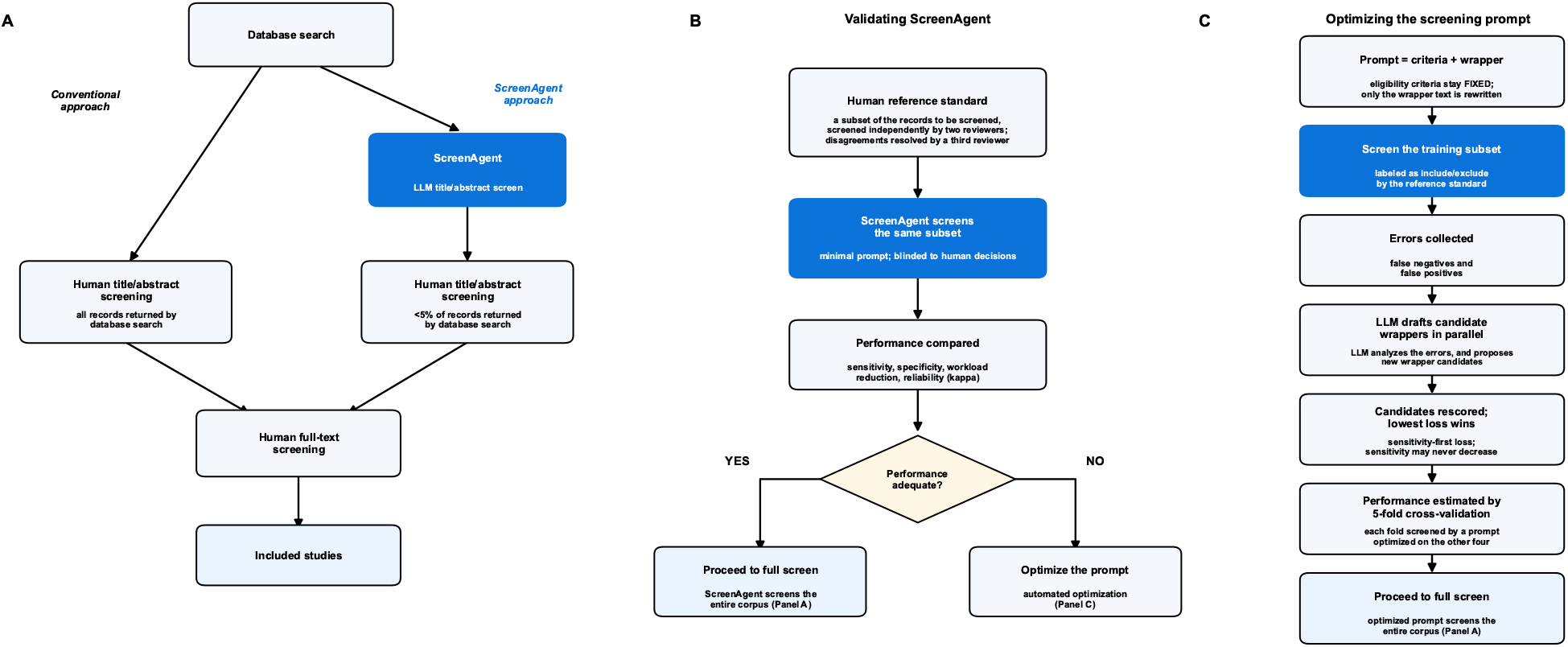
Two-track ScreenAgent pipeline. A broad literature search is screened by the agent; flagged records proceed to human title-and-abstract review and then full-text review, with human reviewers making all final inclusion decisions. The internal review (CBSPS) used a single minimal prompt as-is, whereas the external validation reviews used an automatically optimized, then 5-fold cross-validated, prompt.

### Internal Validation

The prospective application and internal validation were conducted within the CBSPS review, a meta-analysis identifying the magnitude of effect/impact of community-based programs, policies, and laws that reduce suicide. Its central challenge is capturing crossover-effect studies: interventions not primarily designed for suicide prevention (for example, housing support policies, healthcare insurance expansion) that nonetheless affect suicide outcomes, often alongside other health and social outcomes, and are missed by narrow suicide-specific database searches. Internal validation tested whether ScreenAgent recovers these studies from a broad search.

The CBSPS review uses a deliberately broad search (broad suicide and self-harm outcome terms with operators excluding systematic reviews, editorials, and other non-eligible publication types; verbatim strategy in eTable 2)^32^ returning 201,064 unique records across four databases (PubMed, Embase, Web of Science, and PsycINFO), and was deliberately designed to capture both crossover-effect studies and suicide-prevention-specific studies. Its eligibility criteria were developed with each criterion written to be explicit and unambiguous.

The agent screened records with a prompt consisting of a minimal wrapper (the instruction text surrounding the criteria) and the verbatim eligibility criteria (eBox 2) using Opus 4.8^33^ at medium effort. A cascade then re-screened only the agent-included records at maximum reasoning effort, removing false positives at unchanged sensitivity. This cascade is a second, higher-effort pass that re-examines only the records flagged for inclusion by the first stage, tightening specificity without re-screening the full corpus (eTable 6). Because eligible studies are rare in the broad corpus (approximately 1%), a random sample would contain too few includes for robust performance estimates; the test set was therefore drawn using a stricter, more specific search applied to the corpus, enriching for eligible studies (search strategy in eTable 3). As in a conventional systematic review,^34,35^ these 1,000 records were independently double-screened by two reviewers, with disagreements adjudicated to consensus, which identified 44 eligible studies. These 44 became the reference standard, the set of correct includes against which ScreenAgent’s sensitivity was measured. Sensitivity was measured directly against the reference standard on the enriched test set (Figure 2A). Because this enriched set contains a higher share of includes than the broad corpus, workload reduction for the full corpus was estimated separately from a random sample of 1,000 from the full corpus, subsequently confirmed by the full-corpus run. Agreement between ScreenAgent and the adjudicated human consensus was quantified with the Cohen kappa^36^ and percent agreement (Figure 3).

**Figure 2.**
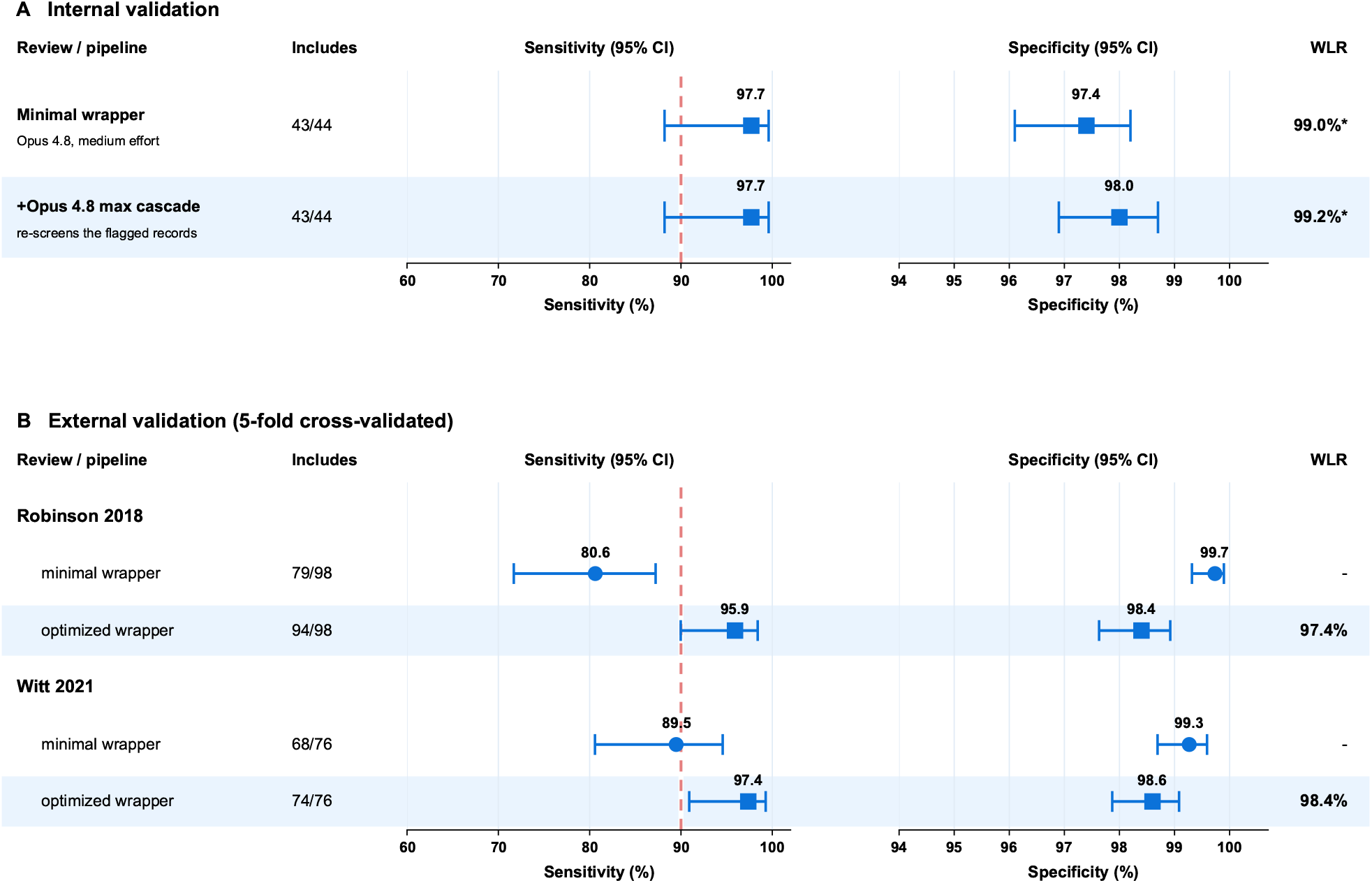
Validation performance. (A) Internal validation on the CBSPS reference standard (1,000 double-screened records; 44 reference-standard includes), showing sensitivity and specificity for the minimal prompt and after the Opus 4.8 maximum-effort cascade. (B) External validation on two published reviews (Robinson 2018; Witt 2021), comparing the minimal prompt with the automatically optimized prompt; sensitivity is the pooled 5-fold cross-validated estimate. Squares indicate point estimates; error bars, 95% CIs; dashed line, 90%.

**Figure 3.**
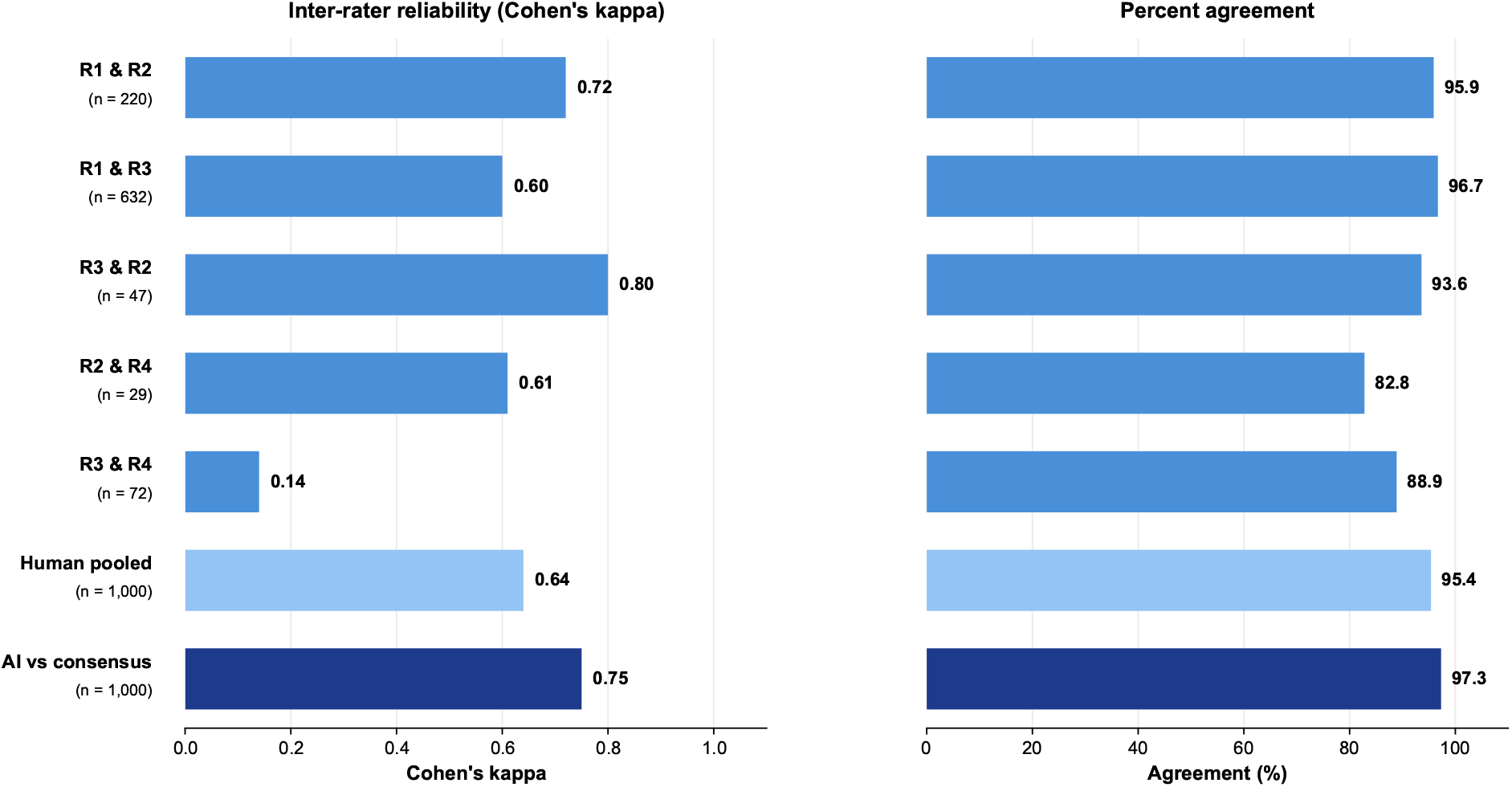
Agent reliability relative to human reviewers.

### External Validation

The external benchmarks were two published systematic reviews with their own eligibility criteria, screening corpora reconstructed from their published search strategies, and ground-truth includes: Robinson comprising 28,267 records with 98 includes and Witt comprising 19,446 records with 76 includes (eTable 4). These reviews were selected because both are recent meta-analyses in suicide prevention with reproducible search strategies and clearly reported eligibility criteria and included studies, giving them the methodological reporting rigor needed to serve as reliable external reference standards. Because the minimal prompt did not achieve acceptable performance, we used an automated procedure that optimizes only the wrapper (eFigure 1). ScreenAgent first screened a training set. Based on the observed errors, five candidate wrappers were proposed in parallel (best-of-5). Each candidate was scored by a sensitivity-favoring loss that penalized false-negative decisions (missed includes) more heavily than false-positive decisions (irrelevant records flagged for inclusion), favoring sensitivity 1.5 times more heavily than specificity, with a squared penalty below a 0.90 performance floor (Figure 4). The candidate with the lowest loss was retained.

**Figure 4.**
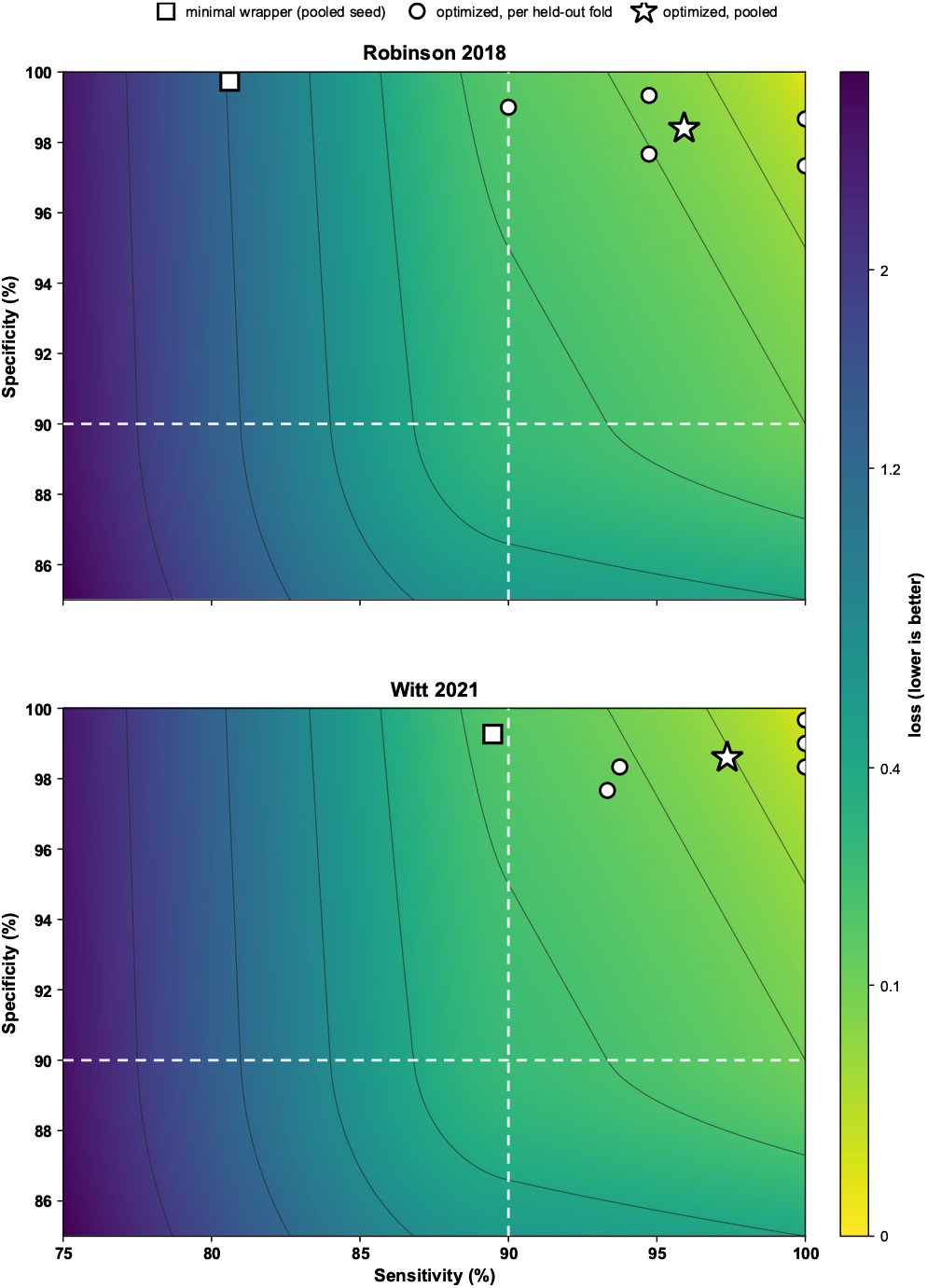
For each external review (Robinson 2018; Witt 2021), the minimal (seed) prompt, the 5 optimized cross-validation folds, and the pooled optimized result are plotted on the sensitivity-favoring optimizer loss function.

Sensitivity was estimated using 5-fold cross-validation^37^ of the ground-truth included studies, divided into five groups. For each fold, the prompt was optimized using the other four groups, then evaluated on the remaining group. Thus, every included study was tested by a prompt that had not been later screened (Figure 2B). This procedure keeps the sensitivity estimate unbiased, with respect to ground-truth includes.

To estimate specificity and workload reduction, a single deployment wrapper trained on all ground-truth positives was run on a prespecified random 1,000-record sample of each reconstructed corpus. Because those positives are contained in its training set, this wrapper was used only for specificity and workload reduction. It was not used to estimate sensitivity, which would have introduced circularity. The complete optimization procedure, including construction of the negative training set, anti-leakage filtering, and the exact loss function, is described in detail in eMethods 2.

### Statistical Analysis

Sensitivity was estimated by direct comparison with the reference standard (internal) and by pooled 5-fold cross-validation (external). Workload reduction and specificity were reported with Wilson 95% confidence intervals^38,39^ (eMethods 2). Statistical analyses were performed in Python, with figures generated using the Matplotlib and NumPy libraries.^40,41^

## Results

### Internal Validation

On the CBSPS testing set, ScreenAgent recovered 43 of the 44 adjudicated eligible studies, yielding a sensitivity of 97.7% (95% CI [88.2%, 99.6%]) using a single minimal, untuned prompt applied verbatim (eBox 2; Figure 2A; Table 1). The single missed study was itself a borderline record for the human reviewers, who disagreed about it at the independent screening stage and resolved it to inclusion only through group adjudication (eBox 1).

**Table 1.** Model and Reasoning-Effort Comparison on the Internal CBSPS Validation Set.

| Model and reasoning effort | Sensitivity, % (95% CI)<br>(No./44) | Specificity, %<br>(single pass) | Specificity, %<br>(cascade) | WLR, %<br>(single pass) | WLR, %<br>(cascade) | Cost per 1000<br>records, \$ |
| --- | --- | --- | --- | --- | --- | --- |
| Opus 4.8, medium | 97.7 (88.2-99.6) (43) | 97.4 | 98.0 | 99.0 | 99.2 | 5.44 |
| GPT-5.5, medium | 97.7 (88.2-99.6) (43) | 98.0 | 98.2 | 99.0 | 99.1 | 6.89 |
| Opus 4.8, no thinking | 95.5 (84.9-98.7) (42) | 97.5 | 98.4 | 99.0 | 99.1 | 5.31 |
| GPT-5.5, no thinking | 93.2 (81.8-97.7) (41) | 97.2 | 98.3 | 98.7 | 99.1 | 3.64 |
| Sonnet 4.6, medium | 86.4 (73.3-93.6) (38) | 98.3 | 98.9 | 98.9 | 99.2 | 2.46 |
| GPT-4.1 | 84.1 (70.6-92.1) (37) | 92.1 | 98.5 | 97.0 | 99.2 | 2.25 |
| Sonnet 4.6, no thinking | 79.5 (65.5-88.8) (35) | 98.0 | 99.0 | 98.7 | 99.2 | 2.45 |
| Haiku 4.5, no thinking | 70.5 (55.8-81.8) (31) | 97.8 | 99.3 | 98.5 | 99.2 | 1.11 |

Abbreviations: CBSPS, Community-Based Suicide Prevention Synthesis; WLR, workload reduction. Sensitivity CIs are Wilson 95% CIs on the 44 reference-standard includes.

The cascade consisted of a second pass using maximum LLM thinking effort that re-screens only the records in the first stage flagged for inclusion. The maximum-effort Opus 4.8 cascade raised specificity from 97.4% to 98.0% with no loss of eligible studies and sensitivity remained at 97.7%. In a comparison on the internal set, the maximum-effort Fable 5 second pass removed about 3 times as many false positives, raising specificity to 98.4% without reduction of sensitivity (eTable 6). This confirms the cascade as a net benefit for workload reduction.

A subsequent full-corpus run excluded 99.4% of all 201,064 records in the single-pass configuration and passing 1,230 for human review. For the cascade step, these 1,230 records were re-screened with a maximum-effort Fable 5 pass; it removed 171 records (13.9%).

### Reliability Relative to Human Reviewers

Agreement between ScreenAgent and the adjudicated human consensus equaled or exceeded the agreement observed between human reviewers (Figure 3). ScreenAgent reached a Cohen kappa of 0.75 against consensus, versus a pooled kappa of 0.64 among human reviewer pairs. Percent agreement showed the same pattern: 97.3% for ScreenAgent versus 95.4% across human reviewer pairs. Human pairwise kappa ranged widely (0.14 to 0.80), reflecting the instability of kappa at low prevalence,^42,43^ which is why percent agreement is reported alongside it. These results place ScreenAgent’s reliability at or above that of trained human screeners.

### External Validation

When applied to two independent published systematic reviews, the minimal wrapper (eBox 2) underperformed, recovering only 80.6% of eligible studies in Robinson 2018 (79 of 98) and 89.5% in Witt 2021 (68 of 76) (Figure 2B; eTable 1). Therefore, we developed an automated procedure to optimize a study-specific wrapper around the fixed eligibility criteria shown in eFigure 1. The procedure generated five candidate wrappers in parallel from the minimal seed prompt and selected the lowest-loss candidate (eFigure 2). In the illustrated Robinson fold, this process increased training sensitivity from 83.3% to 100.0%.

After optimization, pooled 5-fold cross-validated sensitivity on held-out positives rose to 95.9% (95% CI [90.0%, 98.4%]) for Robinson (94 of 98) and 97.4% (95% CI [90.9%, 99.3%]) for Witt (74 of 76). Per-fold sensitivity varied across the held-out folds 90.0% to 100% for Robinson and 93.3% to 100% for Witt (eTable 7 and eTable 8), as expected given the small number of positives per fold.

Figure 4 displays each review’s seed wrapper, held-out folds, and pooled result on the optimizer’s loss surface and shows the nature of the gain. Optimization substantially increased sensitivity while specificity decreased only slightly (Robinson: 99.7% to 98.4%; Witt: 99.3% to 98.6%), reflecting the sensitivity-favoring loss that deliberately trades a small amount of specificity for higher sensitivity. These optimized specificities, together with workload reduction (97.4% for Robinson, 98.4% for Witt), were measured on a prespecified random 1,000-record sample of each reconstructed corpus with a single deployment wrapper (eTable 1), reproducing across both external reviews the internal-validation pattern of high sensitivity with substantial workload reduction.

### Model and Reasoning Effort

To assess whether the internal-validation findings depend on a particular model, we held the minimal prompt fixed and screened the internal sets with eight configurations across five models from two providers (Table 1). Opus 4.8 and GPT-5.5, each at medium effort, tied for the highest sensitivity (both 97.7%, 43 of 44), confirming that the result is not specific to a single model. Medium reasoning effort raised sensitivity over minimum effort (Opus 4.8: 95.5% to 97.7%; GPT-5.5: 93.2% to 97.7%). Smaller models have drastically lower sensitivity (Sonnet 4.6 medium: 86.4%; Haiku 4.5: 70.5%).

### Cost

Screening cost varied by model choice (Table 1). On the internal sets, per-record cost via the Batch API (half the standard rate) ranged from 0.11 cent for Haiku 4.5 to 0.69 cent for GPT-5.5 at medium effort, with the primary Opus medium-to-maximum cascade at 0.54 cent per record. The completed full-corpus screen of all 201,064 records cost $855.91 ($4.26 per 1,000 records). The Fable 5 cascade over these 1,230 records cost an additional $57.70. For the external reviews, the complete optimization runs cost $94.21 for Robinson and $83.71 for Witt (eTable 5). These costs were substantially lower than expected personnel costs of a fully manual title-and-abstract screening at comparable scale.

### Discussion

Suicide is a leading cause of death and a global public health priority,^1^ and the policies and clinical practices intended to prevent it are informed by the evidence assembled in systematic reviews and meta-analyses.^4-7,44^ Those syntheses, in turn, can be no broader than the literature their authors are able to screen. In this study, ScreenAgent identified nearly all eligible studies across three validations, excluded 97-99% of irrelevant records at a fraction of a cent per record, and matched the reliability of trained human reviewers (Cohen kappa 0.75 vs 0.64 for human reviewer pairs). While not designed to replace human judgment, ScreenAgent served as a prescreening tool that made very large literature searches feasible and substantially reduced the manual review burden, addressing a central constraint on the breadth of systematic reviews.

The principal implication is not only faster screening but a change in what reviews can retrieve. Because exhaustive screening of tens of thousands of records is laborious, reviewers have long had to engineer narrow, highly specific search strategies that keep the screening burden manageable. That constraint silently determines which studies can ever enter a review: interventions not primarily targeting the outcome (for suicide, crossover-effect programs such as firearm and alcohol policies, housing support, or healthcare insurance expansion) fall outside a narrow suicide prevention program search and are systematically missed. When automated screening removes this burden, searches can be made deliberately broad, capturing studies a restrictive strategy would otherwise miss. This makes it feasible to pose much broader questions in suicidology, psychiatry, and health policy, capturing the cross-domain evidence that narrow suicide-specific searches cannot reach. For policymakers choosing among prevention strategies, the difference is between evidence limited to expected terminology and evidence wherever it actually appears. Because many of these crossover interventions reduce a range of health and social outcomes beyond suicide, broadening evidence synthesis this way carries relevance and return on investment well beyond suicide prevention alone.

Our findings also argue against simply pointing a capable model at a screening task with generic instructions. A single minimal prompt that performed well on the internal review failed to transfer to reviews with different eligibility criteria, and what restored performance was optimizing how the eligibility criteria were encoded in the prompt, not substituting a more capable model. We therefore propose a general methodology for trustworthy LLM-assisted screening: assemble a small reference set by conventional double screening; use it to optimize and then freeze the screening prompt; estimate sensitivity by cross-validation and estimate specificity and workload reduction on the full corpus, each with confidence intervals; and retain human reviewers as the final decision-makers. This procedure is generalizable, inexpensive relative to manual screening, and converts screening into a measured, reportable component of review.

### Limitations

This study has several limitations. First, every corpus evaluated concerned suicide-related outcomes; although the screening framework is not inherently specific to suicide prevention, its performance in other clinical and policy domains remains to be established. Second, eligible studies were uncommon in each corpus, yielding relatively few true positives and widening the confidence intervals around sensitivity estimates. Third, ScreenAgent’s performance depends on specific language models, whose availability and versions change over time; approaches therefore require re-validation whenever an underlying model changes.^45^ Reported performance should be read as specific to the models and versions evaluated here.

## Conclusions

An LLM screening agent identified nearly all eligible studies with human-level reliability and at low cost, making it feasible to screen literatures that narrow searches systematically miss while keeping human reviewers as the final arbiters. By relaxing the trade-off between search breadth and screening labor, and by making screening performance an empirical, reportable quantity, this approach can strengthen the evidence that guides suicide-prevention policy and clinical practice.

## Data Availability

Relevant data is available from the corresponding author upon reasonable request. Analytic code will be made available upon reasonable request.

## Article Information

### Author Contributions

HCW, DD, PSN, and EEH conceptualized and designed the study. DD developed ScreenAgent and conducted the internal and external validation experiments, with supervision from HCW. AMW, FGS, TR, and DD independently screened titles and abstracts for the internal validation reference standard. DD conducted the statistical analyses. HCW, PSN, EEH, AC, and DD contributed to interpretation of the findings. DD drafted the manuscript and prepared the figures and tables. HCW, PSN, EEH, AC, TR, AMW, DD, and FGS critically revised the manuscript. All authors had full access to all data in the study and had final responsibility for the decision to submit for publication. DD and HCW had direct access to, and verified, all data reported in the manuscript.

