## Supplementary Information for "Making Broad Evidence Synthesis Feasible: An LLM Screening Agent for Meta-Analyses Applied To Suicide Prevention"

**Contents**


- eMethods 1. Screening agent specification
- eMethods 2. Prompt optimization and cross-validation
- eTable 1. Datasets and validation summary
- eTable 2. Broad search strategy (PubMed master string)
- eTable 3. Enriched search strategy (PubMed)
- eTable 4. Systematic reviews used for external validation
- eTable 5. Methodology per external review
- eTable 6. Cascade max-stage results (internal review)
- eTable 7. Per-fold optimizer results, Robinson 2018
- eTable 8. Per-fold optimizer results, Witt 2021
- eTable 9. TRIPOD-LLM checklist
- eFigure 1. Inside the optimizer: 5-fold cross-validation
- eFigure 2. Best-of-5 proposal results
- eBox 1. Crossover-boundary case (the single internal miss)
- eBox 2. Minimal seed prompt (verbatim)
- eReferences


### eMethods 1. Screening agent specification

The screening agent decides, for a single title and abstract, whether a human reviewer should retrieve the full text. For each record it received three inputs: the title, the abstract, and the eligibility criteria for the review being screened. These were assembled into a fixed prompt (the unoptimized form is shown verbatim in eBox 2) and sent to the model.

Rather than emit a bare include or exclude label, the agent returned a structured JSON object with three fields: a decision (include or exclude); a list of *triggered_criteria*, which for an exclude decision names the specific criterion labels the record violates (for example, individual-level treatment, hospital-based program, or outcome not suicide-related); and a one-sentence reason. This feature-extraction design forces the model to justify each exclusion against named criteria rather than make a single holistic judgment, which improves auditability, makes disagreements traceable to a specific rule, and reduces inconsistent borderline calls.

Safety refusals, in which the model declines to process a record on dual-use or content-safety grounds and returns a stop reason of refusal instead of a JSON decision, were treated as confident exclusions, because a legitimate community suicide-prevention program evaluation does not trigger such guardrails; the refusal category was recorded for audit.

### eMethods 2. Prompt optimization and cross-validation

The internal review used a single minimal, untuned prompt. The external reviews used an automated optimizer because that same minimal prompt underperformed on them. The optimizer rewrites only the natural-language wrapper (the instruction text surrounding the criteria); the eligibility criteria and the strict JSON output specification are held fixed throughout.

#### Best-of-5 proposal search

Optimization ran as a single round (round 0). The minimal seed wrapper was first used to screen a training set, and its errors (missed includes and false includes) were collected. A proposer model (Claude Opus 4.8 at high reasoning effort) was then given the seed wrapper and these errors and asked to write five revised candidate wrappers, generated in parallel. This is a best-of-5 search, not an evolutionary or iterative-mutation procedure: the five candidates are independent single-step rewrites of the same seed, and the single best one is selected. There is no mutation across generations.

#### Anti-memorization lint (n-gram check)

Because the proposer model sees the training records when it writes candidates, it could in principle copy paper-specific wording out of a training abstract and into the wrapper. A wrapper that quotes individual training papers would score well on that training set but would not generalize, and would in effect leak the answers into the prompt. To prevent this, every candidate wrapper passed an anti-memorization lint. The wrapper text and each training title and abstract were lowercased and tokenized into words, and any run of six consecutive words (a 6-gram) that appeared verbatim in both the wrapper and any training record was flagged as a memorization violation, which disqualified that candidate. A length of six words was chosen because a shared run that long is almost certainly copied text rather than a coincidental common phrase, giving a signal that is essentially free of false positives. An earlier, stricter version of the lint also screened for proper nouns and specific numbers, but it was removed because it false-flagged generalizable terms (for example, population descriptors such as Indigenous, and standard age thresholds) and starved the optimizer of usable candidates; proper-noun and specific-number screening is now a documented manual reviewer step rather than an automated gate.

#### Recall-weighted loss function

Each linted candidate was scored by a loss that encodes the reviewer priority that missing an eligible study is worse than carrying an extra ineligible one. With $R$ the recall (sensitivity) and $S$ the specificity on the training set, the loss function is:

$$L \;=\; 1.5\,(1-R) \;+\; 100\,\max(0,\;0.90-R)^2 \;+\; 1.0\,(1-S) \;+\; 100\,\max(0,\;0.90-S)^2$$

Above 0.90 on both axes the loss is linear, and a one-point drop in recall costs 1.5 times as much as a one-point drop in specificity, so in the healthy region the optimizer trades specificity for recall at a rate of 1.5 to 1. Below 0.90 on either axis the squared-hinge term (a soft floor) grows steeply and dominates the loss, pulling that axis back above 0.90. These slopes and the 0.90 floors were made to replace any hard accept-or-reject threshold.

#### Acceptance rule

A candidate replaced the incumbent wrapper only if it satisfied three conditions: it did not reduce the number of eligible studies kept (recall did not drop); it produced a net improvement of at least a required number of records, where a single record's worth of improvement is accepted only when it is a recall gain, while a specificity-only edit must correct at least two records (this prevents the optimizer from rewriting the prompt around one idiosyncratic paper); and it achieved a strictly lower loss. Recall is therefore protected at every step.

#### Five-fold cross-validation

To estimate out-of-sample sensitivity without optimizing on the records used to test it, the ground-truth positives were partitioned into five disjoint, size-balanced folds[1] using a fixed random seed. For each fold in turn, the full optimization described above was run using the other four folds (together with the shared in-loop negatives) as the training set, and the resulting wrapper was then applied once to the held-out fold and that fold's held-out negatives. The held-out decisions from all five folds were pooled to produce the reported sensitivity and specificity, with Wilson 95% confidence intervals[2] and an exact paired McNemar test[3] comparing the optimized wrapper with the minimal seed. Because every positive is scored only by a wrapper that never saw it during optimization, the pooled sensitivity is an honest out-of-sample estimate.

#### Negative sampling

Ineligible records (negatives) were drawn into two mutually disjoint pools to prevent any leakage between optimization and testing: a shared in-loop pool of 120 negatives that the optimizer could see while scoring candidates; five held-out pools of 300 negatives each, one paired with each positive fold and used only at test time. The deployment wrapper (below) was trained with the same shared in-loop pool. No negative appears in more than one pool, and the pools were carved deterministically from the negative set by the same seed used for the folds.

#### Deployment wrapper and workload reduction

After cross-validation, a single deployment wrapper was trained on all ground-truth positives at once, using the same best-of-5 procedure, and run over a random sample of 1,000 records from the reconstructed review corpus to measure specificity and workload reduction. Sensitivity is deliberately not reported for this wrapper: because it was trained on the same positives it would then be scored against, its apparent sensitivity would be circular and uninformative. The honest sensitivity estimate is the cross-validated value above, while the deployment wrapper contributes only specificity and workload reduction. All candidate wrappers were capped at 800 words to keep the prompt concise and to limit the room for memorized content.

### eTable 1. Datasets and validation summary

| Review | Topic | Corpus, No. | Includes, No. | Track | Sensitivity, % (95% CI) | Specificity, % | WLR, % |
|---|---|---|---|---|---|---|---|
| Internal (CBSPS) | Community-based suicide prevention (crossover capture) | 201 064 | 44 | Minimal prompt + cascade | 97.7 (88.2-99.6) | 98.0 | 99.2* |
| Robinson 2018 | Youth suicide prevention | 28 267 | 98 | Optimized | 95.9 (90.0-98.4) | 98.4 | 97.4† |
| Witt 2021 | Psychosocial interventions for self-harm (adults) | 19 446 | 76 | Optimized | 97.4 (90.9-99.3) | 98.6 | 98.4† |

Abbreviations: CBSPS, Community-Based Suicide Prevention Synthesis; CI, confidence interval; WLR, workload reduction.
Sensitivity was measured directly against the reference standard for the internal review and by pooled 5-fold cross-validation for the external reviews.
Internal sensitivity and specificity were measured on the enriched 1000-record test set; corpus No. is the full broad-search corpus.
* The completed internal full-corpus run confirmed the estimate: 99.4% WLR across all 201 064 records (single pass). † External WLR was estimated from random 1000-record samples of each corpus (Wilson 95% CI: Robinson 96.2-98.2; Witt 97.4-99.0).

### eTable 2. Broad search strategy (PubMed master string)

Defines the full screening corpus: 201,064 unique records across PubMed, Embase, Web of Science, and PsycInfo after cross-database deduplication (validated PubMed master string shown; translated to the other databases).

**PubMed query**

```
(
suicid* OR "self-harm" OR "self harm" OR "self-harming" OR self-injur* OR "self injur*" OR self-poison*
OR "self poison*" OR parasuicid* OR "self-inflicted" OR "self-directed violence" OR "self-destructive
behavior" OR "self-destructive behaviour" OR "deliberate overdose" OR "intentional overdose" OR
"Suicide"[Mesh] OR "Self-Injurious Behavior"[Mesh]
)
NOT
(
"Editorial"[pt]
OR "Comment"[pt]
OR "Personal Narrative"[pt]
OR "News"[pt]
OR "Newspaper Article"[pt]
OR "Systematic Review"[ti]
)
AND
(
(english[Filter]) AND (2000:2026[pdat])
)
```

### eTable 3. Enriched search strategy (PubMed)

Stricter, more specific search used to draw the 1,000-record internal validation test set.

**PubMed query**

```
(
  "Suicide/prevention"[MeSH]
  OR Suicid*[tiab]
)
AND
(
  "clinical trial"[pt]
  OR "Comparative Study"[pt]
  OR "Evaluation Study"[pt]
  OR "Non-randomized"[tiab]
  OR Nonrandomized[tiab]
  OR cohort[tiab]
  OR observational[tiab]
  OR "Case-control"[tiab]
  OR "cohort studies"[MeSH]
  OR "cross-over studies"[MeSH]
  OR prospectiv*[tiab]
  OR registr*[tiab]
  OR retrospectiv*[tiab]
  OR "propensity score"[tiab]
  OR "Propensity Score"[MeSH]
  OR "Empirical Research"[MeSH]
  OR "Program Evaluation"[MeSH]
  OR laws[tiab]
  OR ecological[tiab]
  OR "interrupted time series"[tiab]
  OR "difference-in-differences"[tiab]
  OR "differences-in-differences"[tiab]
  OR "quasi-experimental"[tiab]
  OR "quasi experimental"[tiab]
  OR "natural experiment"[tiab]
  OR joinpoint[tiab]
  OR "before and after"[tiab]
  OR baseline[tiab]
  OR "pre-intervention"[tiab]
)
AND
(
  program[tiab]
  OR programme[tiab]
  OR prevent*[tiab]
  OR policy[tiab]
  OR policies[tiab]
  OR law[tiab]
  OR legislation[tiab]
  OR regulation[tiab]
  OR "Health Policy"[MeSH]
  OR "Government Programs"[MeSH]
)
AND
(
  communit*[tiab]
  OR populat*[tiab]
  OR nation*[tiab]
  OR "state-level"[tiab]
  OR statewide[tiab]
  OR school[tiab]
  OR schools[tiab]
  OR workplace[tiab]
  OR occupational[tiab]
  OR military[tiab]
  OR "Community Health Services"[MeSH]
  OR "Community Mental Health Services"[MeSH]
)
AND
```

```
(
  "suicide attempt"[tiab]
  OR "suicide attempts"[tiab]
  OR "suicide rate"[tiab]
  OR "suicide rates"[tiab]
  OR "suicide mortality"[tiab]
  OR "suicide death"[tiab]
  OR "suicide deaths"[tiab]
  OR "completed suicide"[tiab]
  OR "suicidal behavior"[tiab]
  OR "suicidal behaviors"[tiab]
  OR "suicidal behaviour"[tiab]
  OR "suicidal behaviours"[tiab]
  OR "Suicide"[MeSH]
  OR "Suicide, Attempted"[MeSH]
)
AND (humans[Filter])
AND (english[Filter])
AND (2000:2026[pdat])
NOT
(
  "Editorial"[pt]
  OR "Comment"[pt]
  OR "Letter"[pt]
  OR "Personal Narrative"[pt]
  OR "News"[pt]
  OR "Newspaper Article"[pt]
  OR "Review"[pt]
  OR "Systematic Review"[pt]
  OR "Meta-Analysis"[pt]
  OR "Published Erratum"[pt]
  OR "Retracted Publication"[pt]
  OR psychotherapy[tiab]
  OR "cognitive behav"[tiab]
  OR "dialectical behavioral therapy"[tiab]
  OR antidepressant[tiab]
  OR pharmacotherapy[tiab]
  OR inpatient[tiab]
  OR "hospital-based"[tiab]
  OR "psychiatric unit"[tiab]
)
```

### eTable 4. Systematic reviews used for external validation

| Review | Topic | Corpus, No. | Includes, No. | Track |
|---|---|---|---|---|
| Robinson 2018 | Systematic review and meta-analysis of interventions designed to reduce suicide-related behavior (suicide, suicide attempts, self-harm, and suicidal ideation) in young people aged 12-25; the 99 included studies spanned clinical settings (52 studies), educational or workplace settings (31 studies), and community-based settings (15 studies). | 28 267 | 98 | Optimized |
| Witt 2021 | Cochrane systematic review and meta-analysis of randomised controlled trials (including cluster-randomised and cross-over designs) evaluating specific psychosocial interventions, delivered after an episode of self-harm, against treatment as usual, enhanced usual care, or an active comparator, in adults with a recent (within 6 months) presentation for self-harm; the primary outcome was repetition of self-harm over up to 2 years of follow-up. | 19 446 | 76 | Optimized |

### eTable 5. Methodology per external review

| Parameter | Robinson 2018 | Witt 2021 |
|---|---|---|
| Topic | Youth suicide prevention | Psychosocial interventions for self-harm (adults) |
| Full corpus | 28 267 | 19 446 |
| Ground-truth includes | 98 | 76 |
| Cross-validation design | 5-fold | 5-fold |
| In-loop negatives | 120 (shared) | 120 (shared) |
| Held-out negatives | 5 x 300 (disjoint) | 5 x 300 (disjoint) |
| Deployment-wrapper training negatives | 120 (in-loop pool) | 120 (in-loop pool) |
| Screening model | Opus 4.8, medium effort | Opus 4.8, medium effort |
| Proposer model | Opus 4.8, high effort | Opus 4.8, high effort |
| Search | Best-of-5 at round 0 | Best-of-5 at round 0 |
| Wrapper length cap | 800 words | 800 words |
| Anti-leakage | n-gram lint + dedup | n-gram lint + dedup |
| Pooled sensitivity, % (95% CI) | 95.9 (90.0-98.4) | 97.4 (90.9-99.3) |
| Pooled specificity, % | 98.4 | 98.6 |
| Cost (USD) | $94.21 | $83.71 |
| Wall-clock | 17.3 min | 14.9 min |

### eTable 6. Cascade max-stage results (internal review)

| Pipeline | Sensitivity, % | WLR, % | FP removed by max | Includes lost |
|---|---|---|---|---|
| Medium only (Opus 4.8) | 97.7 | 93.1 | – | 0 |
| + Opus 4.8 max | 97.7 | 93.5 | 4 | 0 |
| + Fable 5 max | 97.7 | 94.2 | 11 | 0 |

WLR values are measured on the deduplicated internal validation set (n=997; 44 includes, 953 negatives), re-screening the 69 stage-1 flags; Table 1 reports the full 1,000-record test set (68 flagged). Fable 5 removes about 3 times the false positives of Opus-max (11 vs 4) at unchanged sensitivity.

### eTable 7. Per-fold optimizer results, Robinson 2018

| Fold | Selected candidate | Held-out sensitivity, seed % | Held-out sensitivity, optimized % | Held-out specificity, optimized % |
|---|---|---|---|---|
| 1 | Candidate 1 | 80.0 | 90.0 | 99.0 |
| 2 | Candidate 3 | 85.0 | 100.0 | 97.3 |
| 3 | Candidate 2 | 85.0 | 100.0 | 98.7 |
| 4 | Candidate 2 | 78.9 | 94.7 | 97.7 |
| 5 | Candidate 2 | 73.7 | 94.7 | 99.3 |
| Pooled | – | 80.6 | 95.9 | 98.4 |

### eTable 8. Per-fold optimizer results, Witt 2021

| Fold | Selected candidate | Held-out sensitivity, seed % | Held-out sensitivity, optimized % | Held-out specificity, optimized % |
|---|---|---|---|---|
| 1 | Candidate 4 | 81.2 | 93.8 | 98.3 |
| 2 | Candidate 4 | 93.3 | 100.0 | 99.7 |
| 3 | Candidate 2 | 93.3 | 100.0 | 99.0 |
| 4 | Candidate 2 | 86.7 | 93.3 | 97.7 |
| 5 | Candidate 5 | 93.3 | 100.0 | 98.3 |
| Pooled | – | 89.5 | 97.4 | 98.6 |

### eTable 9. TRIPOD-LLM checklist

Completed checklist from Gallifant J, Afshar M, Ameen S, et al. The TRIPOD-LLM reporting guideline for studies using large language models. *Nat Med*. 2025;31(1):60-69. Study type: evaluation of existing LLMs (no de novo development or fine-tuning); LLM task: classification. Page numbers refer to the main manuscript as currently paginated; eTable/eFigure/eMethods/eBox numbers refer to this supplement. Items marked “not stated” or “partial” are open items for the authors.

| Item | TRIPOD-LLM checklist item | Location |
|---|---|---|
| **Title** | | |
| 1 | Identify the study as developing, fine-tuning, and/or evaluating the performance of an LLM, specifying the task, the target population, and the outcome to be predicted. | p1 |
| **Abstract** | | |
| 2 | See TRIPOD-LLM for Abstracts. | pp2-4 |
| **Introduction** | | |
| 3a | Explain the healthcare context / use case and rationale for developing or evaluating the LLM, including references to existing approaches and models. | pp5-7 |
| 3b | Describe the target population and the intended use of the LLM in the context of the care pathway, including its intended users. | p7; pp14-16 |
| 4 | Specify the study objectives, including whether the study describes the initial development, fine-tuning, or validation of an LLM (or multiple stages). | p2; p7 |
| **Methods** | | |
| 5a | Describe the sources of data separately for the training, tuning, and/or evaluation datasets and the rationale. | pp8-10; eTable 1; eTable 2; eTable 4 |
| 5b | Describe the relevant data points and provide a quantitative and qualitative description of their distribution. | eTable 1; eTable 4; eTable 5 |
| 5c | State the date of the oldest and newest item of text used in development and evaluation datasets. | eTable 2 |
| 5d | Describe any data pre-processing and quality checking. | eTable 2; eMethods 2 |
| 5e | Describe how missing and imbalanced data were handled and provide reasons for omitting any data. | p9 (partial) |
| 6a | Report the LLM name, version, and last date of training. | pp13-14; Table 1 |
| 6b | Report details of LLM development process (architecture, training, fine-tuning, alignment). | Not applicable |
| 6c | Report details of how text was generated, including prompt engineering and inference settings. | pp9-10; eBox 2; eMethods 2 |
| 6d | Specify the initial and post-processed output of the LLM. | p9; eMethods 1 |
| 6e | Provide details and rationale for any classification and how thresholds were identified. | p9; eMethods 1 |
| 7a | Include metrics that capture the quality of generative outputs. | Not applicable |
| 7b | Report the outcome metrics' relevance to the downstream task at deployment. | p8 |
| 7c | Clearly define the outcome, how predictions were calculated, the date of inference for closed-source LLMs, and evaluation metrics. | p8; p11 |
| 7d | If outcome assessment requires subjective interpretation, describe the assessors, instructions, and inter-assessor agreement. | p9; Figure 3 |
| 7e | Specify how performance was compared to other LLMs, humans, and benchmarks. | Table 1; Figure 2; Figure 3; p13 |
| **Annotation** | | |
| 8a | If annotation was done, report how text was labeled, including guidelines. | p9 |
| 8b | Report how many annotators labeled the dataset(s), proportion multiply annotated, and inter-annotator agreement. | p12; Figure 3 |
| 8c | Provide information on the background and experience of the annotators. | p17 |
| **Prompting** | | |
| 9a | Provide details on prompt design, curation, and selection. | pp9-10; eMethods 2; eFigure 1; eFigure 2 |
| 9b | Report what data were used to develop the prompts. | pp9-10; eMethods 2 |
| 10 | Summarization preprocessing. | Not applicable |
| 11 | Instruction tuning/alignment details. | Not applicable |
| 12 | Report compute or proxies (time, cost, inference time). | p14; Table 1; eTable 5 |
| 13 | Name the IRB/ethics committee and consent or waiver. | p8 |
| **Open science** | | |
| 14a | Source of funding and role of funders. | p17 |
| 14b | Conflicts of interest and financial disclosures for all authors. | p17 |
| 14c | Indicate where the study protocol can be accessed or state none. | Not stated |
| 14d | Registration information or state not registered. | Not stated |
| 14e | Availability of the study data. | p17 |
| 14f | Availability of the code to reproduce results. | p17 |
| 15 | Patient and public involvement. | Not stated |
| **Results** | | |
| 16a | Flow of text/EHR/patient data through the study. | Figure 1; p12 |
| 16b | Characteristics per data source/setting and split. | eTable 1; eTable 5 |
| 16c | Comparison of clinical variable distributions. | Not applicable |
| 16d | Number of participants and outcome events in each analysis. | pp11-13; eTable 1; eTable 7; eTable 8 |
| 17 | Report LLM performance per prespecified metrics and/or human evaluation. | pp11-13; Figure 2; Table 1 |
| 18 | Report results from any LLM updating. | p12; p14; eTable 6 |
| **Discussion** | | |
| 19a | Overall interpretation including fairness in context of objectives and previous studies. | pp14-16 |
| 19b | Discuss limitations and effects on bias, uncertainty, generalizability. | p16 |
| 19c | Known challenges in using data for the task/domain. | pp5-6; p16 |
| 19d | Define the intended use, input, end-user, level of autonomy/human oversight. | p16 |

| Item | TRIPOD-LLM checklist item | Location |
|---|---|---|
| 19e | How poor quality or unavailable input data should be handled at implementation. | eMethods 1 (partial) |
| 19f | Whether users must interact with input handling or the LLM, and expertise required. | p16 |
| 19g | Next steps for future research, applicability and generalizability. | p16 |

### eFigure 1. Inside the optimizer: 5-fold cross-validation

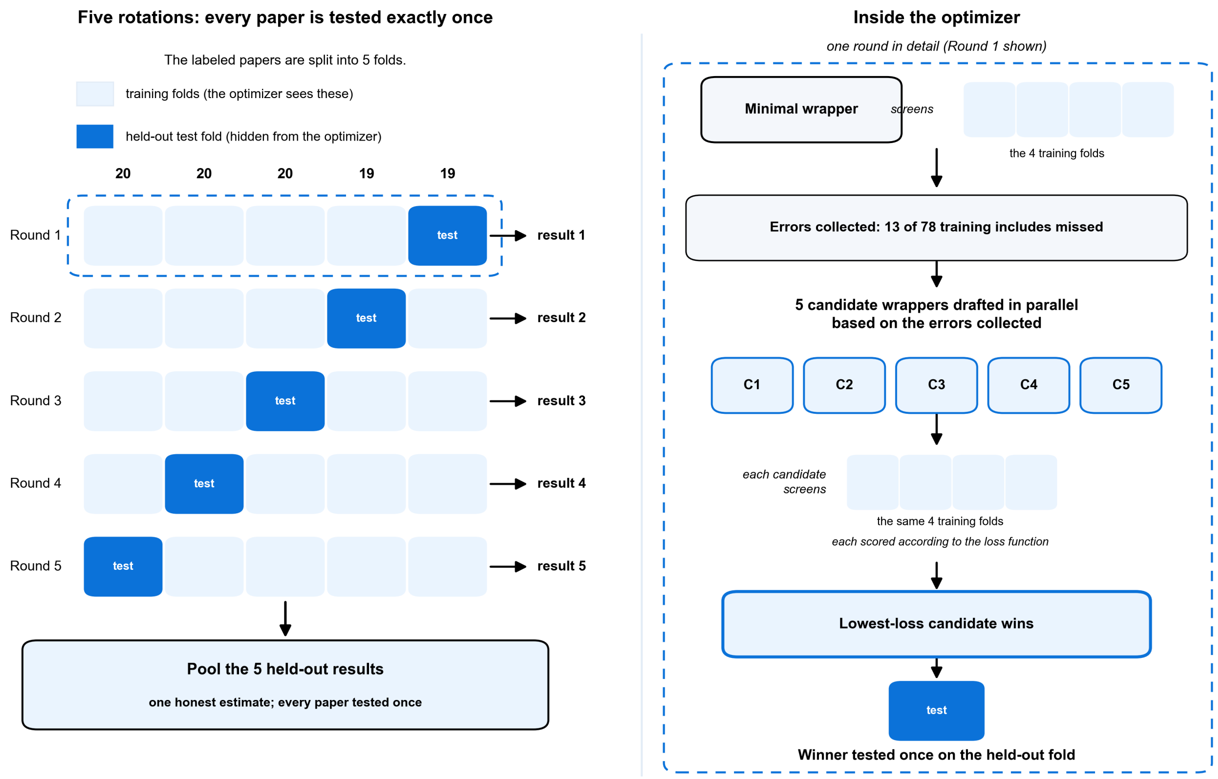


**eFigure 1.** Example numbers show the Robinson 2018 external validation (98 reference-standard includes).

### eFigure 2. Best-of-5 proposal results

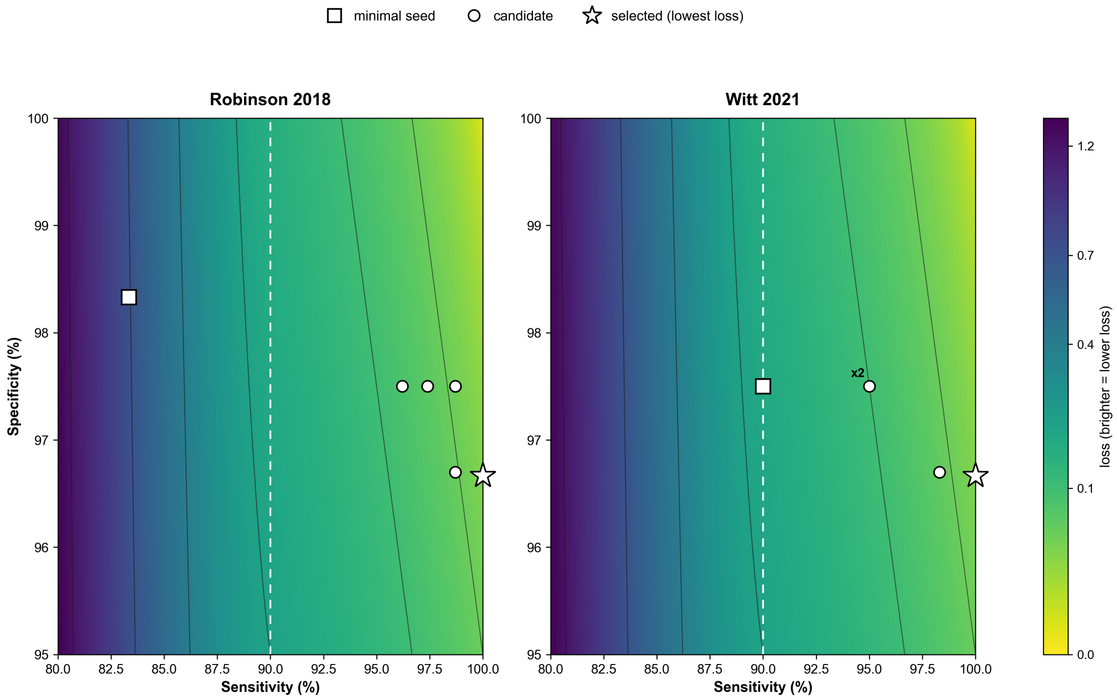


**eFigure 2.** Best-of-5 proposal results are shown for fold 0. Candidates with identical training metrics render as a single point, annotated x2.

### eBox 1. Crossover-boundary case (the single internal miss)

The one internal include the agent missed was "A National Comparison of Suicide Among Medicaid and Non-Medicaid Youth" (Fontanella et al, *American Journal of Preventive Medicine*, 2019). All evaluated models excluded it, judging it a descriptive epidemiological comparison of suicide by population characteristic (Medicaid status) rather than an evaluation of a program or policy effect. This contrasts with policy-effect studies such as analyses of state Medicaid expansion and suicide rates, which are crossover studies and were correctly included. The case sits at the crossover-inclusion boundary and within the human reviewer disagreement band (pooled kappa, 0.64); it is best read as a gold-label boundary call rather than a model failure, and is flagged for adjudication.

### eBox 2. Minimal seed prompt (verbatim)

**System prompt (assembled per review)**

```
You are screening the title and abstract of a study to decide whether a human
reviewer should retrieve the full text for a systematic review. Apply the
eligibility criteria below and decide include or exclude.

Eligibility criteria for this SR:
---
{ELIGIBILITY_CRITERIA}
---

Return STRICT JSON in this exact shape (no extra text, no markdown fences):
{"decision": "include" | "exclude", "triggered_criteria": ["label1", ...], "reason": "one-sentence
explanation"}

For include decisions, triggered_criteria should be [] and reason can be brief.
For exclude decisions, triggered_criteria must list the label(s) from the criteria
that the paper violates.
```

**Per-record user message**

```
Title: {TITLE}

Abstract: {ABSTRACT}
```
